# Effect of gamma sterilization on filtering efficiency of various respiratory facemasks

**DOI:** 10.1101/2020.06.04.20121830

**Authors:** Amit Kumar, D.N. Sangeetha, Ramani Yuvaraj, M. Menaka, V. Subramanian, B. Venkatraman

## Abstract

Three types of respiratory masks viz N95, nonwoven fabric and double layer cotton cloth are being used as an essential inhalation protective measure against COVID-19 by suppressing the entry of respiratory droplets. The filtering efficiency of these masks were tested before and after sterilisation using gamma radiation for the two flow rate conditions corresponding normal breath rate (20lpm) and during sneezing/coughing (90lpm).Sterilisation is carried out using a gamma irradiator containing Co-60 source for the two dose exposures viz. 15kGy and 25kGy.The filtering efficiency for surgical (nonwoven fabric) and double layer cotton cloth mask is found to vary from 18% to 22% for the cumulative particle of size ≥ 0.3µm in both un-irradiated and irradiated condition. The filtration efficiency of N95 mask is found to be reduced to 70% for the most penetrating particle size (0.3 µm) with the flow rate of 20lpm and further reduced for particles in the range of 0.1 and 0.2µm with flow rate of 90 lpm. The reduction in efficiency after gamma sterilization is associated with reduction of electrostatic interaction of filter medium with particles laden in the air stream. Even with reduced filtering efficiency due to gamma sterilisation, the N95 masks are much superior than the surgical and cloth masks. Instead of disposing N95 mask after single use, they can be reused a few times as N70 mask during this pandemic crisis after sterilisation using gamma radiation.

## 1.0 Introduction

One of the essential personal protective equipment (PPE) against COVID-19 infection for the human being is face mask, which offers protection against infectious respiratory droplets or particles like corona virus-laden aerosols around the globe (Feng et al., 2020 and Kin et al.,2020). As millions of masks are being used, it is become necessary for the multiple use of the masks in this pandemic crisis for the public, the staff on the frontline workers in health-care, than others (Avilash et. al., 2020 and Juan et al., 2020). Among the different methods, the penetration levels leading to mask degradation (Juan et al., 2020). Ozone gas appears to be and police force personnel in order to avoid any shortages. It is often considered as good practice to sterilize the masks by killing the secondary pathogens before reusing. Many authorities (governments, manufacturers, scientists and experts) are looking to expand the availability facemask by sterilizing them with effective methods. Under these premises, multiple potential methods for sterilization have begun to be explored. Some are based on chemical methods (hydrogen peroxide, chlorine dioxide, bleach, alcohol, soap solution and ethylene oxide, ozone decontamination etc.) and physical methods (dry /steam heat treatment,UV light sterilization, electron beam and gamma irradiation etc.)(Kumar et al., 2015 and Lei et al., 2020). All above sterilisation method having advantages and disadvantages from one available literature seems to point out that the most promising methods are those that use hydrogen peroxide vapor, ultraviolet radiation, moist heat, dry heat, ozone gas and gamma irradiation. The methods that are not recommended for disinfection or sterilization such as cleaning with soapy water, alcohol, bleach immersion, ethylene oxide, microwave, high temperature, autoclave or steam because they can significantly degrade the filter, because they alter or degrade the electrostatic properties of the filter fibre sand affect particle effective in decontaminating respirators mask without damaging the facemask, although it presents risks for the safety and health of workers who carry out the process if it is not handled properly (Zhang et al., 2004 and Juan et al., 2020). Gamma irradiation is safe, reliable and highly effective at treating a wide variety of products with varying densities.

Radiation sterilization is used since late 1950s to eliminate microorganisms, such as bacteria, fungi and spores, from medical equipment. Currently, almost 50 per cent of healthcare gamma rays, electron-beams or X-rays prior to their use. With the ability to penetrate and distribution process by facilitating final packaged products as well as raw material needs, ensuring full sterility of the product.

It is found in some of the recent work on the Covid-19 pandemic scenario to determine whether radiation could be used for sterilization of facemasks (Avilash et. al., 2020, Tzu et surgical mask) and self-made two-ply cotton masks are tested for filtering efficiency with and work, testing in accordance with two breath condition (normal and during sneezing) is Three types of facemasks viz. N95, non-woven fabric (surgical mask) and self-made double layer cotton cloths have been tested for particulate filtering efficiency for the two flow rates products, such as gloves, syringes and single use protective clothing, are sterilized using products while sealed in their final packaging, gamma irradiation supports the manufacturing al., 2020, Man et al., 2020 and Lei et al., 2020). In this context, three types of masks, which are being used most of the countries, include N95, non-woven fabric masks (often called as without gamma sterilisation. The comparison of filtering efficiency of the current work and with the results available in literature is made for N95 masks, in particular, in the present highlighted. As millions of masks are being used daily, in order to prevent the spread of secondary pathogens, cross contamination and continue to use the irradiated N95 face mask after sterilisation at par with surgical and cloth masks, filtering efficiency tests were conducted before and after sterilisation for the three types of masks and the results are described in this paper.

## 2.0 Material and methods

Three types of facemasks viz. N95, non-woven fabric (surgical mask) and self-made double cotton cloths have been tested for particulate filtering efficiency for the two flow rates viz., 20 and 90 lpm condition corresponding to normal breathing rate and during the sneezing/coughing respectively (Lee et al., 2019). The tests were conducted using the filter test rig at HEPA Filter Testing Laboratory, Radiological and Environmental Safety Division (Fig.1). It consist of a cylindrical tube of 8 cm diameter and 180 cm length connected with sampling port in the upstream and downstream. Provision is available for fixing the facemask and to measure the pressure drop across the facemask. The airflow rates (20 and 90lpm) were achieved through air displacement pump and air flow meter. K type thermocouple is used for the measurement of temperature of air stream. Two numbers of Optical Particle Counter (OPC)(Model and Make: 1.108 and M/s Grimm Aerosol Technik, Germany),one for upstream and other for downstream, are used for particle counting from the size range 0.3µm to 20µm. Sequential Mobility Particle Spectrometer (SMPS)(Model and Make: 5.416 and M/s Grimm Aerosol Technik, Germany)is used for the particle counting from the size range of 20nm to 1.1µm. Aerosol Generator (Model and Make: 7.811 and M/s Grimm Aerosol Technik, Germany) is used for the generation of test aerosols. The polystyrene latex (PSL) particle sizes 1.0 ± 0.1µm (M/s MAGSPHERE Inc., USA) and 102.7 ± 1.3 nm (M/s Polysceince Inc., USA) are used as mono dispersed test aerosols.

**Fig. 1.**
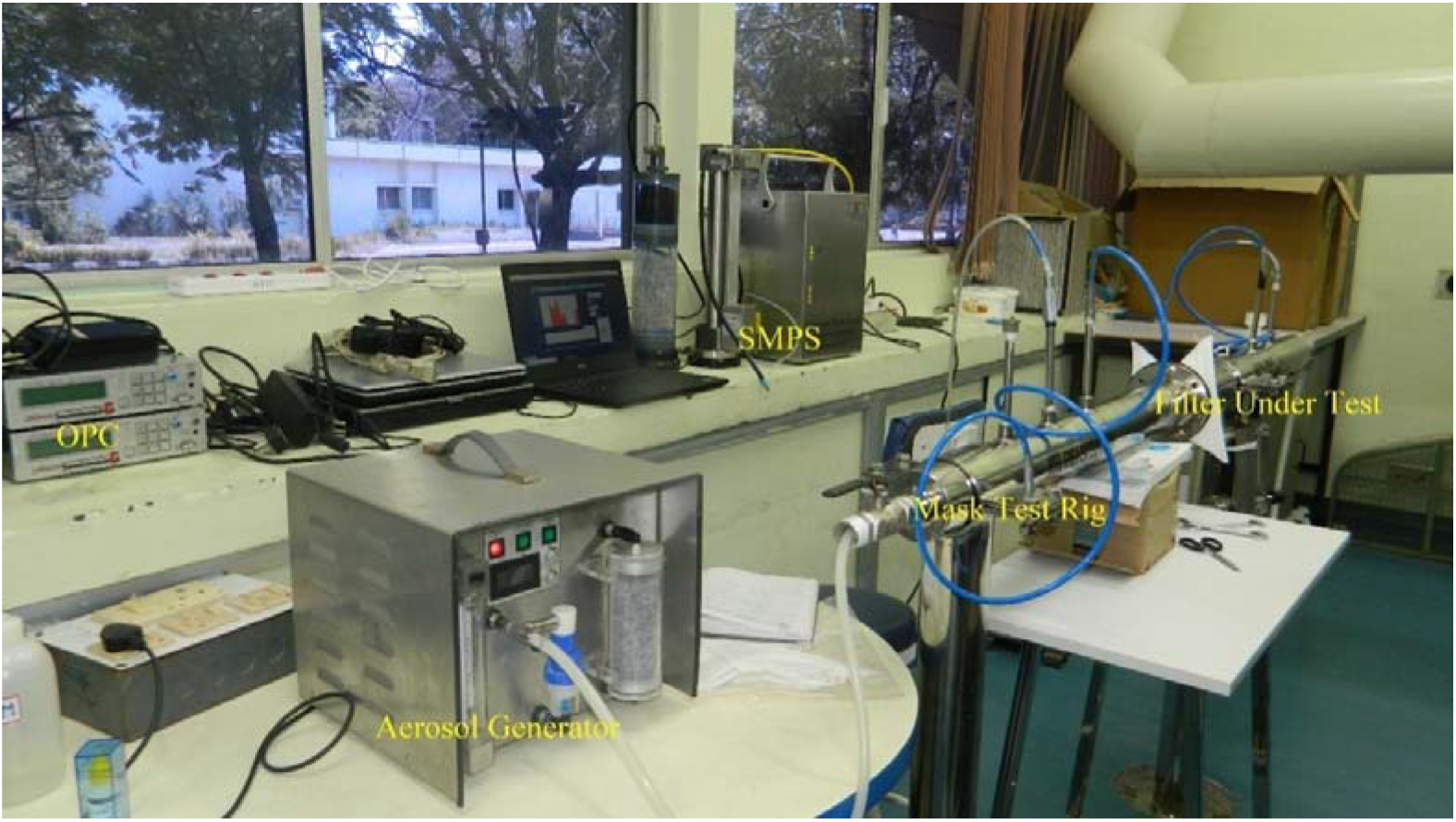
Photograph of the experimental setup.

The sterilization of various mask is achieved by using Gamma Chamber (Model and Make: GC500 and M/s BRIT, India) where the masks were irradiated using Co-60 source for the desired dose level. A set of three N95 facemask and 10 numbers of each of surgical and cloth masks were irradiated to doses of 15kGy and 25kGy. It is to be noted that the recommended dose for medical product sterilisation and other organic compounds ranges from 15–25kGy (Techdoc, BRIT, 2015).The number of mask tested in each type are summarised in Table 1. The image of nonwoven fabric, double layer cotton cloth and N95 facemask are shown in Fig. 2.

**Table 1.** Number and type of mask tested for two exposure doses.

| Type of facemask | Gamma radiation dose |  |  |
| --- | --- | --- | --- |
|  | 0 (kGy) | 15 (kGy) | 25(kGy) |
| **Surgical Mask(Nos.)** | 20 | 10 | 10 |
| **Cloth Mask(Nos.)** | 20 | 10 | 10 |
| **N95 Mask (Nos.)** | 6 | 3 | 3 |

**Note: 0 kGy corresponds to control pieces (un-irradiated).**

**Fig. 2.**
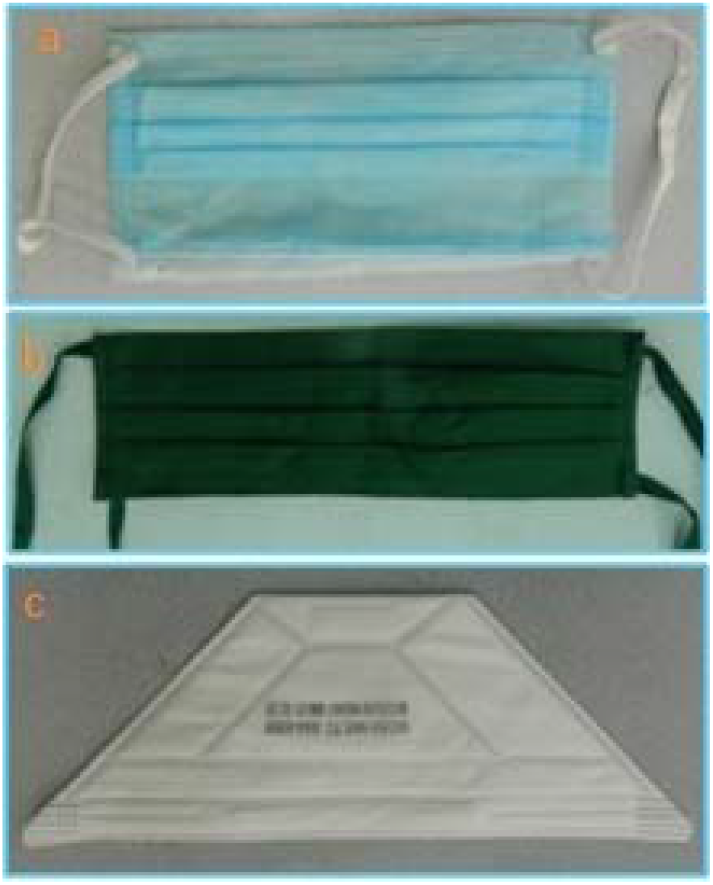
Image of (a)nonwoven fabric,(b) double layer cotton cloth and (c) N95 facemask.

The masks were inserted into a specialized air duct and ambient particulates were blown through the filter in the duct with a face velocity of 0.4 to 0.5 m/s and 2.5 to 2.6 m/s corresponding to 20±0.2 lpm and 90±1 lpm flow rates respectively. The ambient air temperature is measured as 26 ±1°C and RH% is 65 ±3. In the next run, mono dispersed aerosols of 102.7 nm and 1.0 µm were generated and filter testing is carried out for the same two flow rate conditions.

## 3.0 Results and discussion

### 3.1 Testing with ambient aerosols size ≥0.3µm

The filtration efficiency of masks is determined for atmospheric aerosols of size ≥ 0.3µm and is shown in Fig. 3.The pressure drop across the filter is measured and is given in Table 2. The pressure drop indicates condition for usage during breathing and is found to be in the accepted range (inhalation and exhalation resistance limit is 35 and 25 mm w.g. respectively). It is observed from Fig. 3 that particulate filtering efficiency is less for high flow rate (90 lpm) when compared to the low flow rate (20 lpm) for all three types of mask.

**Fig. 3.**
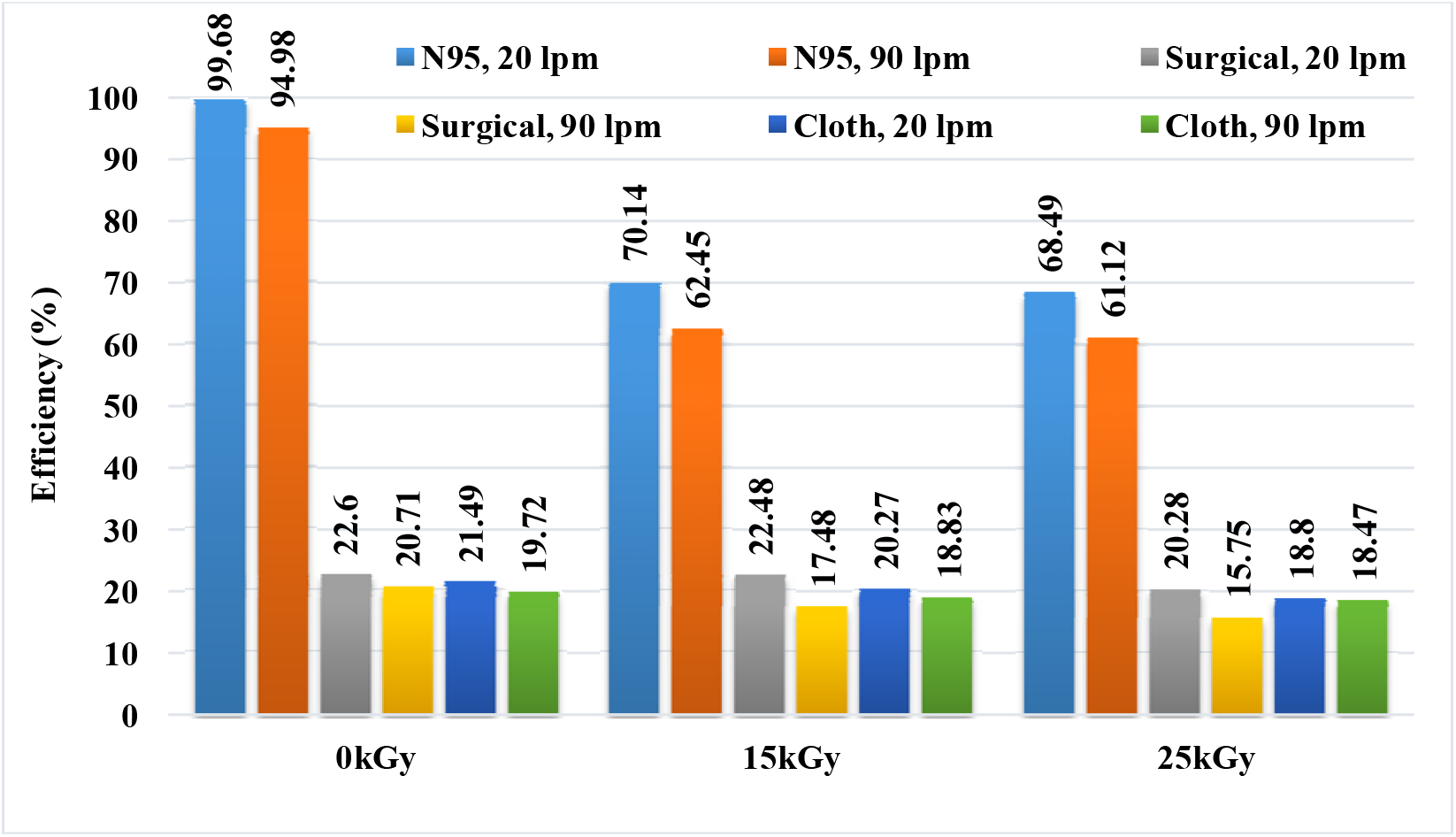
Filtration Efficiency of mask for atmospheric aerosols of size ≥0.3µm

**Table 2.** Pressure drop across the filter for two-flow rate condition.

| S. N. | Type of Mask | Flow rate (lpm) | $\Delta P$ (mm of water gauge) |
| --- | --- | --- | --- |
| 1. | N95 | 20 | 3.15 |
|  |  | 90 | 26 |
| 2. | Surgical | 20 | 0.25 |
|  |  | 90 | 0.75 |
| 3. | Cloth | 20 | 1.18 |
|  |  | 90 | 7.5 |

The surgical and cloth mask are performed more or less same or slightly decrease in efficiency after sterilisation when compared to without gamma sterilized condition. The filtering efficiency found to vary from 18% to 22% in both un-irradiated and irradiated condition for the cumulative particles of size ≥0.3µm.In"0.3µm.In the case of N95 masks, the filtering efficiency is found to be reduced to 70% from 99.6% and about 62% from 95% for the two irradiated conditions for 20 lpm and 90 lpm respectively. Further, it is also noticed that there exists about 1–2% reduction in efficiency for all the three types of masks between 15 kGy and 25 kGy exposed doses.

### 3.2 Testing with aerosol size 1.0µm

Fig. 4 shows the filtering efficiency of these three types of filters for mono dispersed PSL particles of size 1.0µm with and without sterilisation and for the two flow rate conditions. The filtering efficiency for N95 mask shows>96%even after sterilization for both the flow rate conditions. The filtering efficiency of surgical mask is found to be in the range of 70–83% before sterilisation and reduced to 43–56% after gamma sterilisation. The filtering efficiency of cloth mask showed 55–63% before sterilisation and 45–59% after sterilisation. The cotton mask showed not much variation after sterilisation. Upon comparison with filtering efficiency of surgical and cloth masks between the particles in the size ≥0.3µm and mono dispersed particles of size 1.0 µm, it is observed that filtering efficiency for the surgical and cloth masks is found to be more for 1.0µm particles for both flow rates under sterilized or non-sterilized condition. It is to be noted that, that cumulative efficiency for particles ≥0.3µm is shown in Fig. 3 and about 95% of the particles are in the range of 0.3 and 0.5 µm in the ambient aerosols for which efficiency is about 20%, whereas, this test is carried out with mono dispersed particles of 1.0µm, for which the efficiency is found to be higher. It is also to be noted that the filtration efficiency of all three types of mask is more for 90 lpm flow rate for 1.0 µm sized particles when compared to the 20 lpm. This is due to higher filtration of micron size aerosols by impaction at large flow rate condition.

**Fig. 4.**
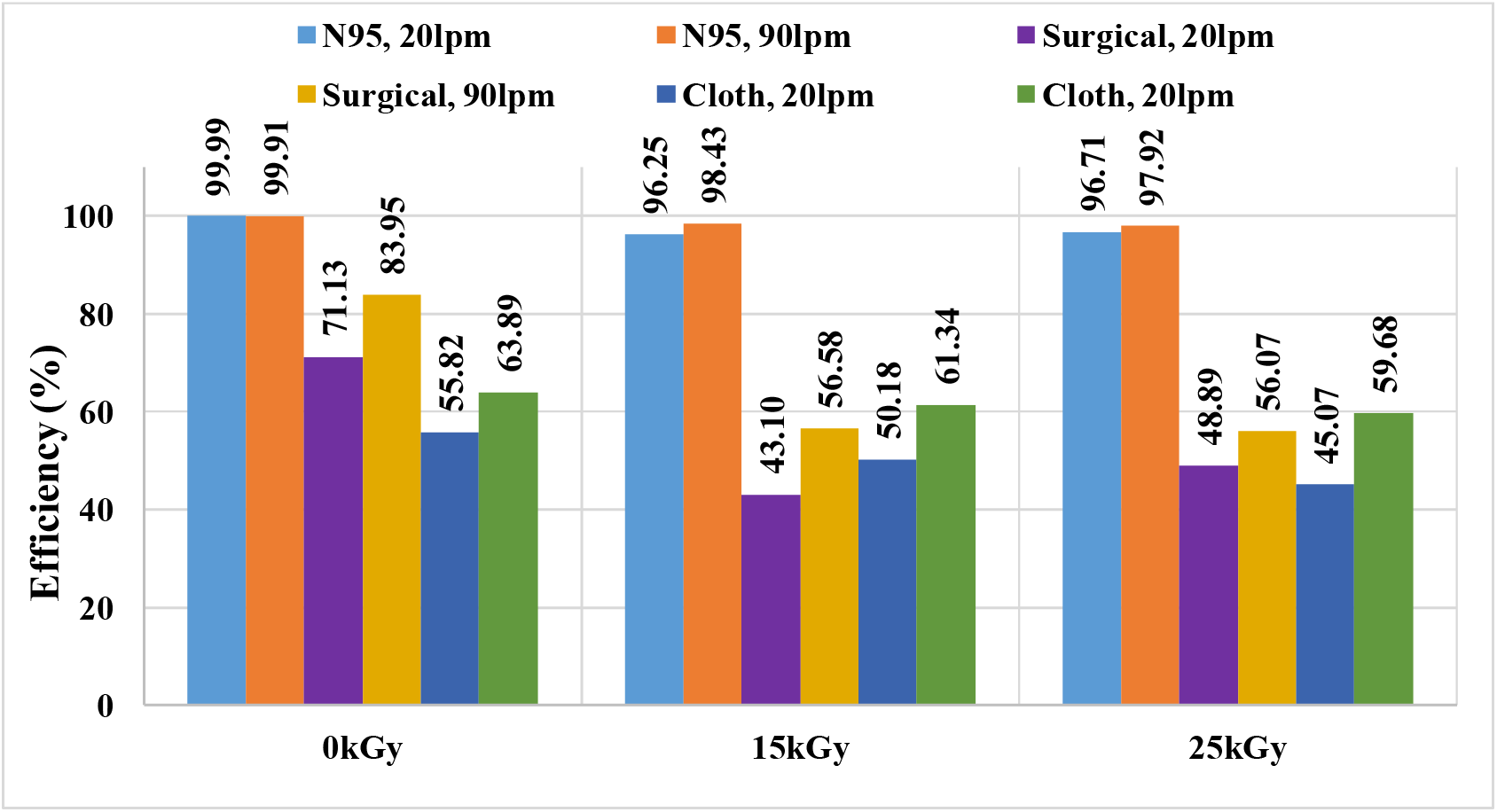
Mask Efficiency with irradiation and two flow rate for 1.0 µm PSL aerosols.

### 3.3 Testing with aerosol size 100nm for N95 mask

The N95 facemask is exclusively tested for102.7nm laboratory generated PSL aerosols in line with size range of viruses (70–100 nm) (Kim et al., 2020).Fig. 5 shows filtration efficiency of N95 mask for both un-irradiated and gamma sterilized and for two flow rate conditions. It can be seen from the figure that, the filtering efficiency is more than 95% for un-irradiated condition and for both flow rates. The efficiency found to be reduced from 99% to 94% for the flow rate of 20lpm and 95% to 72% for 90lpm under gamma-sterilized condition. It can be seen from Fig. 4 that, the filtration efficiency of N95 mask is decreased for 90 lpm flow rate when compared to the 20 lpm for gamma sterilisation conditions. This is opposite to1.0 µm aerosols where the particles are carried away due to high flow rate by the flow gas streamlines in the filtering media.

**Fig. 5.**
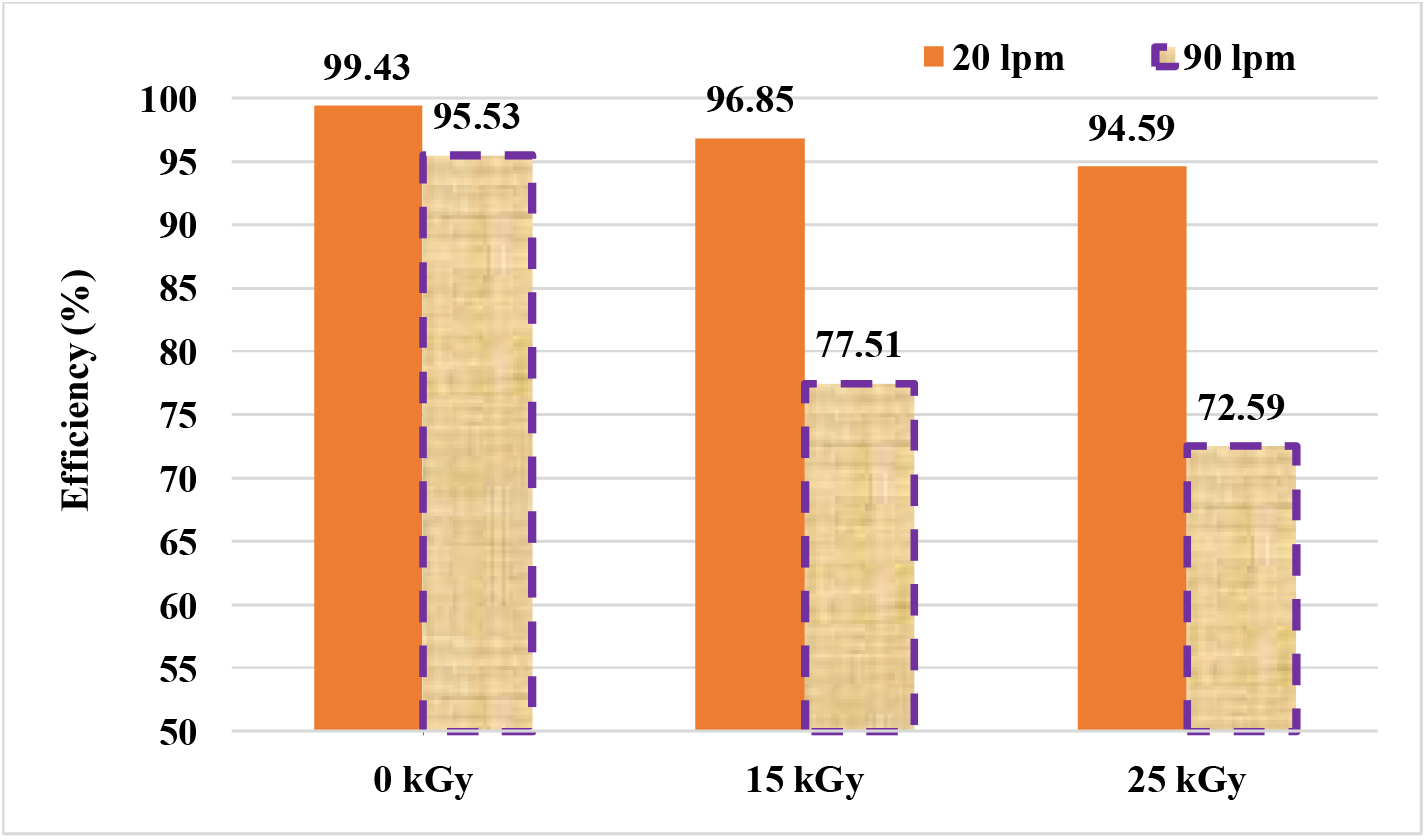
N95 facemask filtering efficiency for 102.7 nm PSL aerosols.

### 3.4 Filtering Efficiency for N95 masks for particles from 25nm to 5.0µm

The filtering efficiency of N95 mask is examined by combining data of OPC and SMPS covering from the range of 25nm to 5 µm sized aerosols, because, coughs and sneezing actions produce droplets < 100 nm (< 0.1 µm) and bulk of droplets > 100 nm (>0.1 µm) (Lee et al., 2019).The particle generated from coughing/sneezing ranges from 0.1 µm to 100 µm (Yang et al., 2007, Gralton et al., 2011 and Lindsley et al., 2012).The filtration efficiency of N95 mask under all conditions (un-irradiated, gamma sterilised and for two flow rates) is shown in Fig. 6. It is observed from Fig. 6 that, the filtering efficiency under un-irradiated condition is greater than 99% for the all the particles except in the range of 0.2–0.3 µm where it is reduced by about 3–8%. Under the gamma sterilized condition the filtration efficiency falls down to about 70% for aerosols in the size range of 0.2–0.3µm for 20 lpm and it is reducing further for high flow rate (90 lpm).

**Fig. 6.**
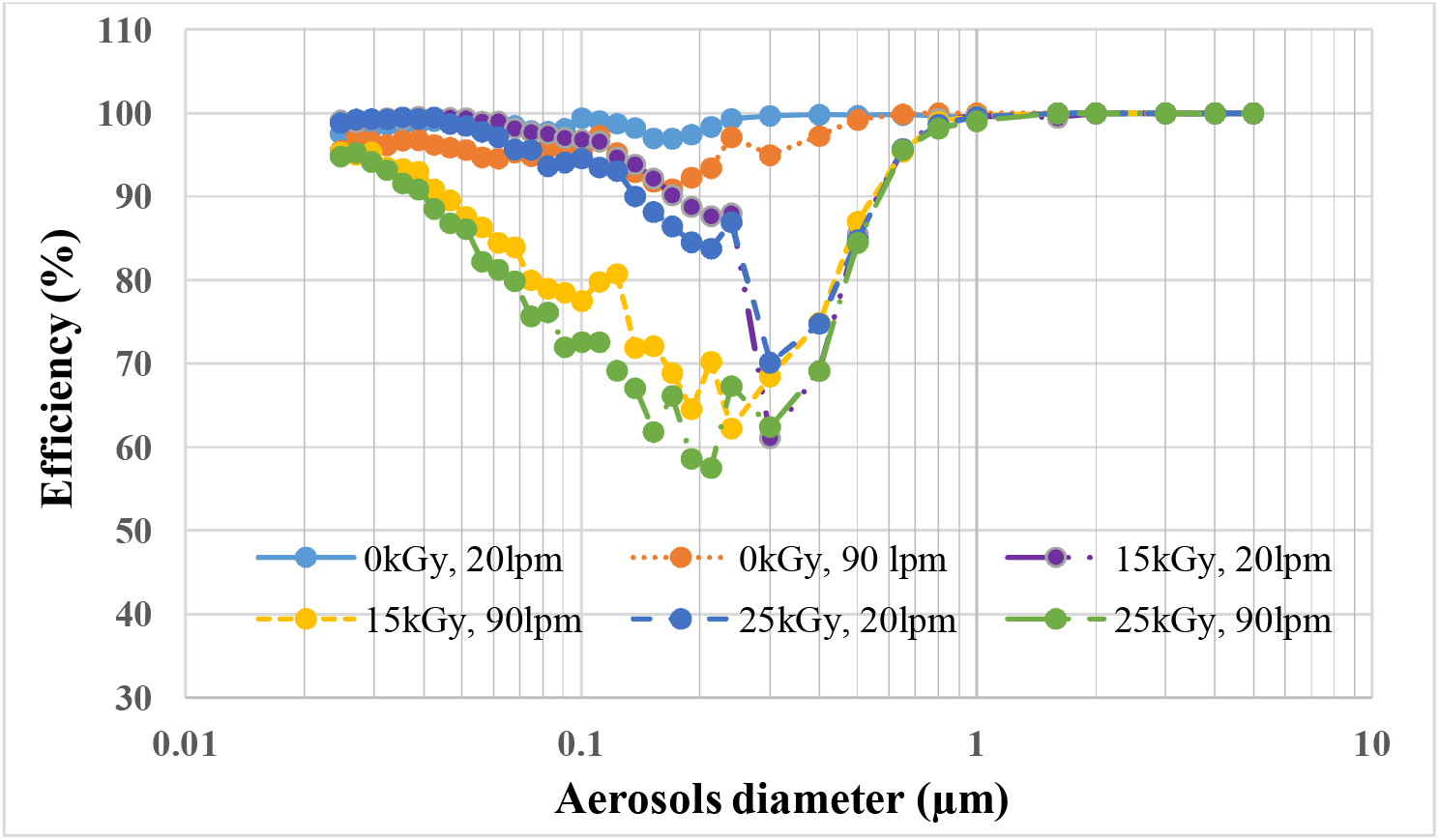
Efficiency of N95 mask for the aerosols of size ranging from 25nm to 5µm, for 15kGy and 25kGy dose exposure and for the two flow rates 20lpm and 90lpm.

It is known that N95 filter consists of electrostatic filtration media which encompass a broad class of materials that are capable of capturing and retaining fine air borne particulates through electrostatic interaction (Coulomb and dielectrophoretic forces) in addition to mechanical processes (Impaction, settling, Interception and Diffusion) (David L. Myers and B. Dean Arnold, 2003). The reduction in efficiency due to radiation sterilization is attributed as follows: Upon irradiation, the static charges associated with fibrous filters become neutralized (David L. Myers and B. Dean Arnold, 2003).It is known that, least efficiency is associated with particles in the range of 0.2-0.3µm that is bigger for diffusion and smaller for interception; hence, the efficiency is increased for this range by electrostatic interaction. When the media lost its charges, the particles are captured only by mechanical process where the efficiency is reduced from 99% to 70%. The fall of efficiency confirmed from Fig. 3 and Fig. 5, in the case of 1.0µm particle, the efficiency is not found reduced much i.e. 99.9% to 97% even for gamma irradiation.

### 3.5 Comparison of N95 mask filtering efficiency from literature

A comparison of filtering efficiency of N95 respiratory mask with various works from the literature under gamma-sterilized condition is made and summarized in table 3. It can be seen from table that, the irradiation dose for mask sterilization is varied from 10 to 50kGy and mask efficiency has been tested for aerosols ranging from 0.1 to 1.0µm. The reduction in efficiency is found to be more for most penetrating particle size (0.3 µm) in all the cases. Among all the works carried out, the work of Avilash et al., 2020 showed highest reduction in efficiency under gamma sterilized condition. This is may be due to their filtering media as control mask itself is showing 5–15% less efficiency from others works. Further, the filtering characteristics of mask much depends on fibre diameter, packing density, charge on fibres and filter thickness, as well as aerosol characteristics like diameter and density of aerosols and airflow velocity (flow rate). The filtering efficiency of present work is more comparable with Man et al., 2020, but here also, at what flow rate that masks have been tested is not given. It is noted that in all the works, the testing flow rates has not been mentioned. However, it is important to test the masks under breathing rate condition towards fit for the purpose.

**Table 3.**
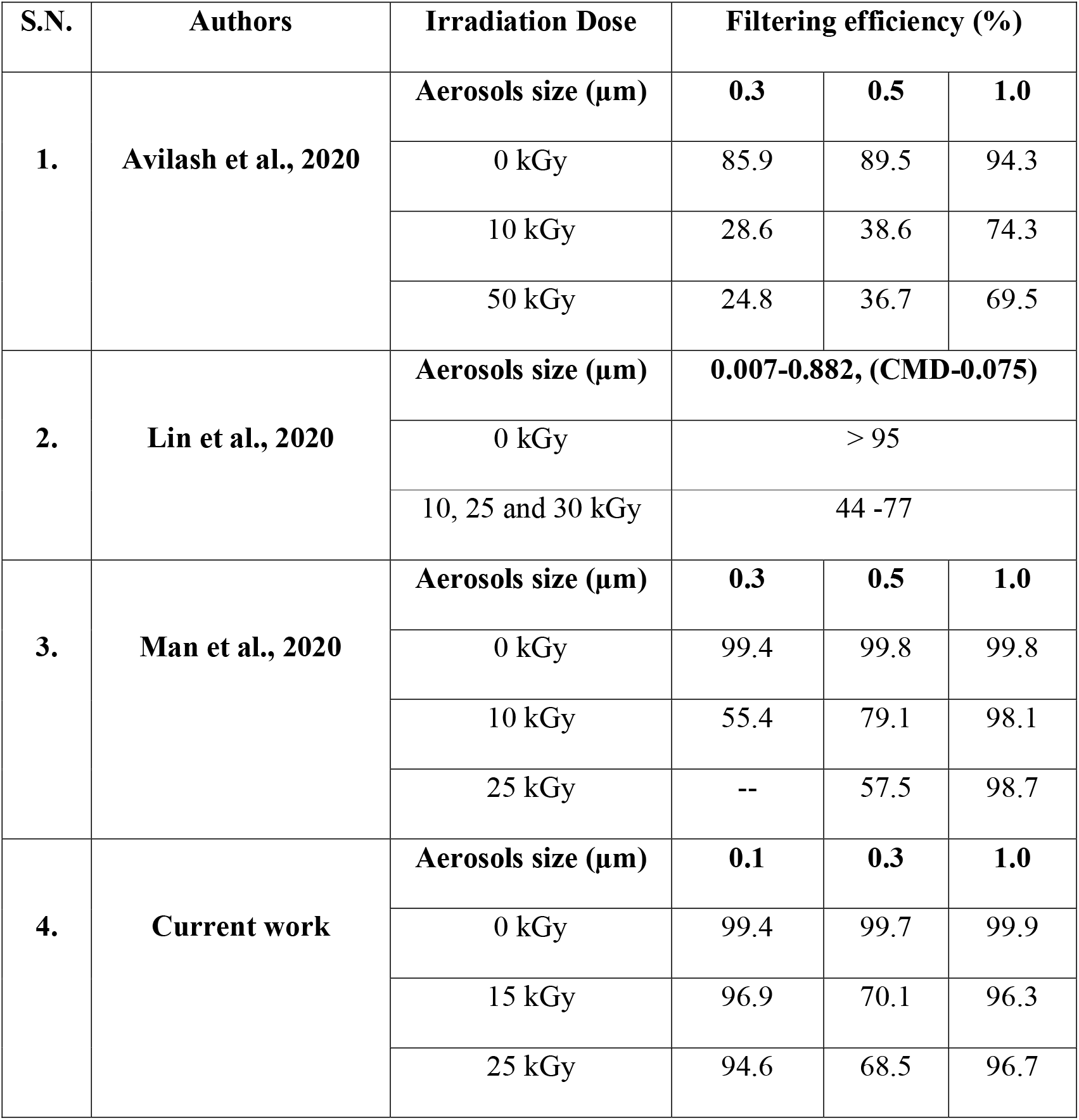
Comparison of N95 respiratory mask filtering efficiency from literature.

## 4.0 Summary and Conclusion

Various types of facemask viz. N95, surgical mask and self-made double layer cloth mask have been tested for particulate filtering efficiency for two-flow rate condition viz. 20 and 90 lpm before and after sterilization using gamma radiation. The sterilization was carried out for two-dose condition viz. 15kGy and 25kGy.The masks were tested using ambient aerosols and using mono dispersed polystyrene latex particles of size 1.0 µm and in particular,N95 mask is tested with 102.7 nm particles.

The filtering efficiency for surgical and cloth mask is found to vary from 18% to 22% for the cumulative particle of size ≥0.3µm in both un-irradiated and irradiated condition. The lower efficiency is attributed to density of fibres in the non-woven type masks while the density of wrap and weft per unit area in the case of woven cloths. However, these masks are effective for the particles in the size range of 1.0 µm and above (about 50% efficiency). It is also noticed that, filtering efficiency is found to be reduced by only 1–2% after sterilization for the both surgical and cloths masks for the ambient aerosols sized ≥0.3µm.

In the case of N95 mask, the gamma sterilization has shown decrease in efficiency from 99% to about 70% and still lesser with higher flow rate for ambient aerosols. The reduction in efficiency of N95 mask after irradiation may be attributed to neutralization charges in the electrostatic filters and in particular, only in the size range of 0.2–0.3µm where mechanical filtration is only effective and electrostatic interaction is ineffective. The filtration efficiency is found to be more than 70% for particles lesser than 0.1 µm and greater than 0.5 µm. Further, filtration efficiency increases more than 90% with decrease of particles size less than 0.1µm and 99.9% for particles greater than 0.5µm.

Various masks showed no significant changes in fit or measurable structural changes when exposed to the 25kGy dose. The breathability test (pressure drop) and filtering efficiency conveys that, the double or triple layer of cotton mask could be potential substitute for medical or surgical mask for respiratory infected person in sterile environment. The healthy or common population may daily use cotton mask in the social community since it is washable and reusable. Since, the particle generated from coughing/sneezing is ranging from 0.1µm to 100 µm (Yang et al., 2007, Gralton et al., 2011 and Lindsley et al., 2012), the surgical and cotton masks significantly reduce (more than 50%) the microorganism expelled during coughing/sneezing. In the populated country like India and to avoid any shortage condition of disposable surgical masks, the common self-made cotton mask may be potential option for healthy people. Further, even with reduced filtering efficiency, the N95 masks after gamma sterilisation is found to be better option than the surgical and cloth masks and can be recommended for volunteers, police personnel and health workers in case of shortage. Instead of throwing or disposing N95 mask after single use, they still could be reused as N70 mask during this pandemic crises after sterilisation using gamma radiation.

## Data Availability

Yes

## 5.0 Acknowledgement

The authors acknowledge to the Dr. C. V. Srinivas, Head, RIAS and Dr. R. Venkatesan, Head, RESD for his encouragement and support to carrying out this work.

## Notes

### Competing Interest Statement

The authors have declared no competing interest.

